# Open-source ImageJ analysis of fundus autofluorescence in early dry age-related macular degeneration: gray-level intensity by pattern and interobserver agreement of pattern grading

**DOI:** 10.64898/2026.07.18.26358399

**Authors:** Jesús Noel Jaurrieta-Hinojos, Ángeles Yahel Hernández-Vázquez, Gerardo González-Saldívar, Adriana Saucedo-Castillo, Alejandro Babayán-Sosa, Juan Abel Ramírez-Estudillo

## Abstract

**Background:** Fundus autofluorescence (FAF) patterns in early age-related macular degeneration (AMD) are usually graded qualitatively, and quantitative analysis depends on proprietary calibrated systems. We evaluated open-source ImageJ for gray-level FAF analysis and interobserver agreement of the Bindewald classification.

**Research design and methods:** Retrospective single-center study of FAF images (2010–2016) of eyes with early dry AMD. Mean, minimum, and maximum 8-bit gray levels were measured in a 300-pixel circular region centered on the fovea. Two graders independently classified each image; agreement was assessed with Cohen’s kappa.

**Results:** Forty-five eyes of 32 patients (mean age 74.2 ± 8.5 years; 62.5% women) were included. Mean gray level was 120.3 ± 24.1. Seven of eight patterns were identified, most often minimal change (31.1%). Mean gray level ranged from 88.4 (lacelike) to 143.8 (reticular), without a significant difference among patterns (p = 0.146). Graders agreed on pattern in 28 eyes (62.2%; κ = 0.53; 95% CI, 0.33–0.71). Agreement was fair for foveal involvement (κ = 0.32) and lesion extent (κ = 0.34).

**Conclusions:** ImageJ allows simple gray-level FAF measurement, but the values are uncalibrated and not comparable with quantitative autofluorescence. Interobserver agreement for Bindewald patterns was moderate, lower than in the original description of the classification.

## 1. Introduction

Age-related macular degeneration (AMD) is a leading cause of irreversible visual impairment in people older than 65 years. Fundus autofluorescence (FAF) is a noninvasive imaging modality that maps the fluorophores of the retinal pigment epithelium (RPE), mainly lipofuscin, whose accumulation and loss are central to AMD pathogenesis [1].

Bindewald et al. described eight FAF patterns in early AMD: normal, minimal change, focal increased, patchy, linear, lacelike, reticular, and speckled [2]. The same group described additional patterns in the junctional zone of geographic atrophy [3], which were later related to atrophy progression [4,5]. These classifications are qualitative. In the original description, two graders agreed in 82% of 100 eyes (κ = 0.77; 95% CI, 0.67–0.87) [2], but agreement outside the group that developed the classification has seldom been reported.

Quantitative autofluorescence (qAF) addresses the limitations of visual grading by referencing the signal to an internal fluorescent standard [6]. It requires dedicated hardware, a specific acquisition protocol, and proprietary software, so it cannot be applied to the large archives of conventional FAF images that many centers hold. ImageJ (National Institutes of Health, Bethesda, MD, USA) is an open-source image analysis program that reads common image formats and reports gray-level statistics for user-defined regions of interest (ROI) [7,8].

This two-phase retrospective study had two aims. The first was to describe a simple ImageJ protocol for gray-level measurement of archived FAF images in early dry AMD and to report gray-level intensity across Bindewald patterns. The second was to measure interobserver agreement of Bindewald pattern grading, lesion extent, and foveal involvement between two independent graders.

## 2 Patients and methods

### 2.1. Study design and setting

This single-center, retrospective study of consecutive cases was conducted at the Department of Retina, Fundación Hospital “Nuestra Señora de la Luz,” I.A.P., Mexico City, Mexico. The protocol was approved by the Institutional Ethics and Research Committees of Fundación Hospital “Nuestra Señora de la Luz,” I.A.P.. Because the study was retrospective and noninterventional, informed consent was not required. Patient data were handled confidentially and in accordance with the Declaration of Helsinki.

### 2.2. Patients and images

The electronic records were searched for all patients who underwent FAF imaging with a combined confocal scanning laser ophthalmoscope and optical coherence tomography device (Spectralis HRA+OCT; Heidelberg Engineering, Heidelberg, Germany) between January 2010 and September 30, 2016. Images were retrieved as 768 × 768-pixel files (JPG, TIF, or PNG), and the diagnosis of each case was reviewed.

Eyes with early dry AMD were included. Exclusion criteria were poor image quality, duplicate images, incorrect diagnosis, neovascular AMD or geographic atrophy in the evaluated image, and ocular comorbidity that could confound FAF interpretation. Both eyes of a patient were included when both were eligible. Eyes with advanced AMD were catalogued separately and are not analyzed here.

### 2.3. Image analysis (phase 1)

Images were analyzed with ImageJ version 2.0.0-rc-54/1.51h (Fiji distribution) using the following steps (Figure 1):

1. Open the image file in ImageJ.
2. Set the spatial scale (Analyze > Set Scale) to 95 pixels/mm.
3. Convert the image to 8-bit grayscale (Image > Type > 8-bit), giving a gray-level range of 0–255.
4. Adjust display contrast and apply pseudo-color lookup tables as needed for visual inspection. These settings change the display only and were not applied to the pixel data.
5. Place a circular ROI 300 pixels in diameter (3.16 mm; area 7.83 mm^2^) centered on the fovea.
6. Record the mean, minimum, and maximum gray level within the ROI (Analyze > Measure).
7. Export the results to a spreadsheet and save an annotated copy of the image.

**Figure 1.**
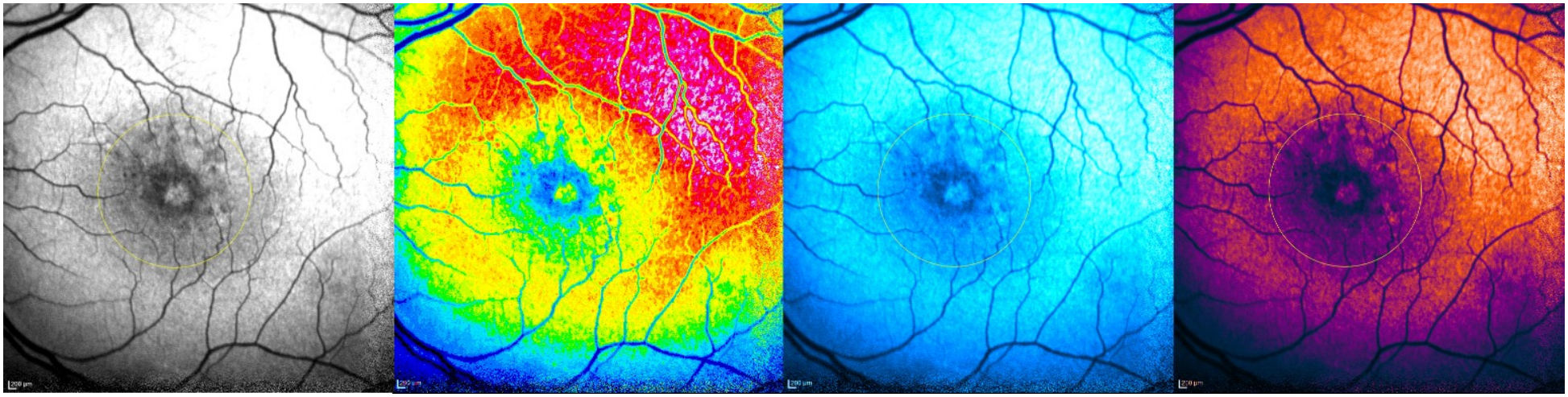
A fundus autofluorescence image displayed in ImageJ with four lookup tables: grayscale (A), Fire (B), Ice (C), and Orange Hot (D). The circle marks the 300-pixel region of interest centered on the fovea. Lookup tables change the display for visual inspection and do not alter the 8-bit gray levels used for measurement.

Gray-level measurements were made once, by the first grader. The number of discrete lesions within the ROI was also recorded.

### 2.4. Pattern grading and interobserver agreement (phase 2)

Two graders independently classified each image according to the Bindewald classification [2]. For each eye they also recorded lesion extent (focal or diffuse; not applicable when no lesion was present) and foveal involvement (present or absent). Each grader was masked to the other’s results. After independent grading was complete, discordant cases were reviewed with a third, senior grader. Because adjudicated labels are not independent, they were not used to compute agreement.

### 2.5. Statistical analysis

Continuous variables are reported as mean ± standard deviation (SD) and range, and categorical variables as counts and percentages. Patient-level variables (age and sex) are reported per patient; image variables are reported per eye. Gray level was compared across patterns, as graded by the first grader, with the Kruskal–Wallis test. This comparison is exploratory: several patterns had three or fewer eyes, and the test does not account for the inclusion of both eyes of some patients. Agreement between fellow eyes was summarized with a one-way intraclass correlation coefficient.

Interobserver agreement was assessed with percent agreement and unweighted Cohen’s kappa (κ), because the patterns are nominal categories. Ninety-five percent confidence intervals (CI) were obtained by bootstrap with 5000 resamples, resampling patients rather than eyes to account for the correlation between fellow eyes. Kappa values were interpreted according to Landis and Koch [9]: 0.21–0.40 fair, 0.41–0.60 moderate, 0.61–0.80 substantial, and 0.81–1.00 almost perfect. Analyses were performed with Python 3 (NumPy, SciPy).

All agreement statistics in this version were computed on the independent, pre-adjudication gradings. Versions 1 and 2 of this preprint reported higher kappa values that had been computed after adjudication of discordant cases; those values are superseded by the present analysis.

## 3. Results

### 3.1. Study population

Approximately 800 FAF studies were identified, and the records of 423 patients were reviewed. AMD was present in 139 patients: 107 (77.0%) with dry AMD and 32 (23.0%) with neovascular AMD in one or both eyes. Among patients with dry AMD, 15 images were excluded for poor quality. Of the 92 evaluable images, 45 (48.9%) showed early AMD and 47 (51.1%) showed advanced AMD.

The early AMD cohort comprised 45 eyes of 32 patients; 13 patients contributed both eyes. Twenty patients (62.5%) were women. Mean age was 74.2 ± 8.5 years (range, 60–94). Twenty-three right eyes and 22 left eyes were included. The mean number of discrete lesions per eye was

2.2 ± 3.4 (range, 0–12). Cohort characteristics and gray-level measurements are shown in Table 1.

**Table 1.** Characteristics of the cohort and gray-level measurements.

| **Variable** | **Value** | **Range** |
|---|---|---|
| Patients, n | 32 |  |
| Eyes, n | 45 |  |
| Patients contributing both eyes, n | 13 |  |
| Age, years (per patient), mean ± SD | 74.2 ± 8.5 | 60–94 |
| Women, n (%) | 20 (62.5) |  |
| Right / left eyes, n | 23 / 22 |  |
| Lesions per eye, mean ± SD | 2.2 ± 3.4 | 0–12 |
| Mean gray level, mean ± SD | 120.3 ± 24.1 | 74.8–160.8 |
| Minimum gray level, mean ± SD | 32.1 ± 13.5 | 3–63 |
| Maximum gray level, mean ± SD | 205.8 ± 30.7 | 125–255 |

SD: standard deviation. Gray levels are uncalibrated 8-bit values (0–255) measured in a 300-pixel circular region of interest centered on the fovea.

### 3.2. Gray-level measurements and pattern distribution

Mean gray level within the ROI was 120.3 ± 24.1 (range, 74.8–160.8). The mean of the minimum values was 32.1 ± 13.5 (range, 3–63), and the mean of the maximum values was 205.8 ± 30.7 (range, 125–255).

According to the first grader, seven of the eight Bindewald patterns were present; no eye showed the speckled pattern. The most frequent pattern was minimal change (14 eyes, 31.1%), followed by focal increased (11, 24.4%), patchy (7, 15.6%), normal (5, 11.1%), lacelike and reticular (3 each, 6.7%), and linear (2, 4.4%). The fovea was involved in 13 eyes (28.9%). Lesion extent was focal in 22 eyes and diffuse in 19; it was not applicable in 4 eyes graded as normal.

Mean gray level by pattern is shown in Table 2 and Figure 2. It was highest in eyes with the reticular pattern (143.8 ± 16.1; n = 3) and lowest in eyes with the lacelike pattern (88.4 ± 18.5; n = 3). The ranges of individual eyes overlapped widely between patterns, and the difference among patterns was not statistically significant (Kruskal–Wallis H = 9.53, 6 degrees of freedom; p = 0.146).

**Table 2.** Gray-level measurements and lesion characteristics by fundus autofluorescence pattern (first grader; n = 45 eyes).

| **Pattern** | **Eyes, n** | **Mean gray level, mean ± SD** | **Minimum, mean** | **Maximum, mean** | **Lesions, mean** | **Foveal, n** | **Diffuse / focal, n** | **Extent NA, n** |
|---|---|---|---|---|---|---|---|---|
| Normal | 5 | 128.5 ± 15.1 | 30.4 | 208.8 | 0.0 | 0 | 0 / 1 | 4 |
| Minimal change | 14 | 118.4 ± 26.3 | 35.6 | 196.4 | 0.1 | 1 | 9 / 5 | 0 |
| Focal increased | 11 | 119.5 ± 21.8 | 32.0 | 205.7 | 3.8 | 4 | 2 / 9 | 0 |
| Patchy | 7 | 128.2 ± 19.1 | 29.6 | 220.4 | 6.7 | 4 | 5 / 2 | 0 |
| Linear | 2 | 101.3 ± 35.3 | 27.0 | 180.5 | 1.5 | 2 | 0 / 2 | 0 |
| Lacelike | 3 | 88.4 ± 18.5 | 17.7 | 189.3 | 1.7 | 1 | 0 / 3 | 0 |
| Reticular | 3 | 143.8 ± 16.1 | 42.3 | 244.0 | 0.0 | 1 | 3 / 0 | 0 |
| Speckled | 0 | – | – | – | – | – | – | – |
| All eyes | 45 | 120.3 ± 24.1 | 32.1 | 205.8 | 2.2 | 13 | 19 / 22 | 4 |

SD: standard deviation; NA: not applicable (no lesion recorded). Gray levels are uncalibrated 8-bit values (0–255). Kruskal–Wallis test for mean gray level across the seven observed patterns: p = 0.146.

**Figure 2.**
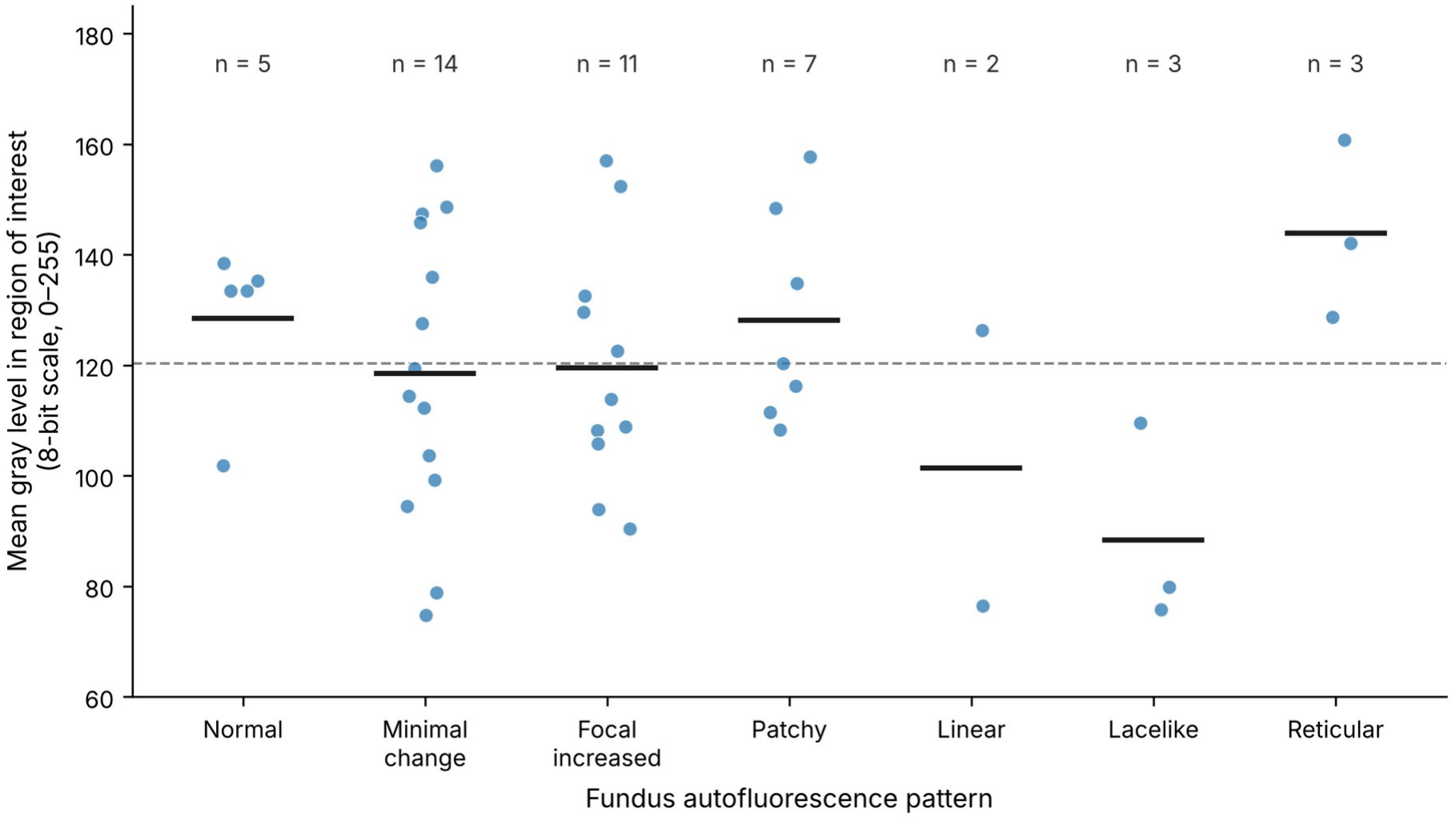
Mean gray level within the region of interest by fundus autofluorescence pattern, as graded by the first grader (n = 45 eyes). Each dot is one eye; horizontal bars show the mean for each pattern; the dashed line shows the overall mean (120.3). Gray levels are uncalibrated 8-bit values.

In the 13 patients who contributed both eyes, mean gray level was correlated between fellow eyes (intraclass correlation coefficient, 0.65; mean absolute difference, 16.0 gray levels), although the two eyes were assigned the same pattern in only 7 patients.

### 3.3. Interobserver agreement

All 45 eyes were graded independently by both graders (Table 3). The graders assigned the same Bindewald pattern in 28 eyes (62.2%), corresponding to moderate agreement (κ = 0.53; 95% CI, 0.33–0.71). The cross-classification is shown in Table 4. The most frequent disagreement was between the normal and minimal change patterns (5 of 17 discordant eyes). The remaining disagreements involved the focal increased pattern versus the lacelike, patchy, minimal change, or reticular patterns (8 eyes), the reticular pattern versus the minimal change or patchy patterns (3 eyes), and the patchy versus the lacelike pattern (1 eye).

**Table 3.** Interobserver agreement between two independent graders (pre-adjudication gradings).

| **Variable** | **Eyes, n** | **Agreement, n (%)** | **κ** | **95% CI** | **Interpretation** |
|---|---|---|---|---|---|
| Bindewald pattern | 45 | 28 (62.2) | 0.53 | 0.33–0.71 | Moderate |
| Foveal involvement | 45 | 29 (64.4) | 0.32 | 0.09–0.55 | Fair |
| Lesion extent (focal / diffuse) | 41 | 27 (65.9) | 0.34 | 0.05–0.63 | Fair |

κ: unweighted Cohen’s kappa; CI: confidence interval, by bootstrap with patients as the resampling unit (5000 resamples). Interpretation according to Landis and Koch [9]. Lesion extent was analyzed in eyes rated by both graders.

**Table 4.** Cross-classification of Bindewald patterns by the two graders (n = 45 eyes).

| **Grader 1 \ Grader 2** | **Normal** | **Minimal change** | **Focal increased** | **Patchy** | **Linear** | **Lacelike** | **Reticular** | **Total** |
|---|---|---|---|---|---|---|---|---|
| Normal | 1 | 4 | 0 | 0 | 0 | 0 | 0 | 5 |
| Minimal change | 1 | 9 | 2 | 0 | 0 | 0 | 2 | 14 |
| Focal increased | 0 | 0 | 7 | 2 | 0 | 2 | 0 | 11 |
| Patchy | 0 | 0 | 0 | 5 | 0 | 1 | 1 | 7 |
| Linear | 0 | 0 | 0 | 0 | 2 | 0 | 0 | 2 |
| Lacelike | 0 | 0 | 1 | 0 | 0 | 2 | 0 | 3 |
| Reticular | 0 | 0 | 1 | 0 | 0 | 0 | 2 | 3 |
| Total | 2 | 13 | 11 | 7 | 2 | 5 | 5 | 45 |

Rows: first grader; columns: second grader. Concordant gradings lie on the diagonal (28 eyes).

Agreement on foveal involvement was fair (29 of 45 eyes, 64.4%; κ = 0.32; 95% CI, 0.09–0.55). The second grader recorded foveal involvement in 25 eyes and the first grader in 13. Agreement on lesion extent was also fair (27 of 41 eyes with a rating from both graders, 65.9%; κ = 0.34; 95% CI, 0.05–0.63).

After review of the discordant cases with the third grader, three eyes could not be assigned a consensus pattern.

## 4. Discussion

This study had two findings. First, gray-level statistics can be obtained from archived FAF images with a short, fully open-source protocol. Second, when two graders classified the same early AMD images independently, agreement on the Bindewald pattern was moderate, and agreement on foveal involvement and lesion extent was only fair.

The gray-level values reported here are not quantitative autofluorescence. Calibrated qAF references the signal to an internal fluorescent standard and corrects for acquisition settings and ocular media, which makes values comparable between eyes, sessions, and centers [6,10].

Conventional FAF images are acquired with operator-adjusted sensitivity and are commonly normalized by the device software, and gray level is further affected by lens opacity and image compression. An 8-bit gray level therefore describes the brightness of a given exported image and cannot be read as lipofuscin content. Two observations in our data agree with this interpretation. Gray level did not differ significantly among patterns, and individual values overlapped widely. In addition, gray level was correlated between the fellow eyes of the same patient even when the two eyes showed different patterns, which suggests that patient-level and acquisition-level factors account for a substantial part of the variation.

For the same reason, the ranking of patterns by mean gray level should not be interpreted. The reticular pattern had the highest mean, but this rests on three eyes, and calibrated studies have generally found that qAF in early and intermediate AMD is not increased, and is often reduced, relative to age-matched controls [6,11–13]. In the study by Gliem et al., none of 40 patients with early or intermediate AMD had qAF values above the 95% prediction interval of 108 controls [6]. Our data neither support nor contradict a prognostic meaning for pattern-specific intensity.

Within these limits, the protocol has practical uses. ImageJ is free, runs on any operating system, and reads images exported from any device, so the same steps can be scripted as a macro and applied to an entire archive. Gray-level statistics from a fixed ROI may be useful for within-image descriptions, such as the contrast between a lesion and its surroundings, and as a first step toward standardized handling of legacy FAF images in centers without access to calibrated systems. Any use for comparison between eyes or over time would require calibration or validation against qAF.

Interobserver agreement for the Bindewald pattern (κ = 0.53) was lower than the κ of 0.77 reported by the authors of the classification, who agreed in 82% of 100 eyes [2]. Several factors may explain the difference. The original graders developed the classification, whereas ours were not involved in its development and applied it from the published description. Our sample was small, and several patterns were rare, which makes kappa unstable. The patterns are also not mutually exclusive in practice: an eye can show features of more than one, and the grader must choose the predominant one. The disagreements we observed were concentrated at boundaries where this choice is hardest, between normal and minimal change and between focal increased and the patchy or lacelike patterns.

Agreement was lower still for foveal involvement and lesion extent, and the two graders differed systematically: one recorded foveal involvement in almost twice as many eyes as the other.

These variables were not defined in detail in our protocol, and the fovea cannot be located precisely on FAF alone because macular pigment attenuates the central signal. Explicit definitions, reference images, and co-registration with optical coherence tomography would be expected to improve agreement, and we consider them necessary before these variables are used in research.

These results have a bearing on automated analysis. Deep learning classifiers of FAF patterns are trained and evaluated against human labels. If independent human graders agree at a moderate level, single-grader labels are a weak reference standard, and reported classifier accuracy should be read against measured human agreement, with adjudicated or multi-grader labels used for training.

This study has limitations. It was retrospective, from a single center, and small, with three or fewer eyes for three of the seven patterns observed. Gray level was uncalibrated and measured on exported image files of mixed formats. Images were acquired between 2010 and 2016 and were not paired with optical coherence tomography, so patterns could not be related to structural findings such as subretinal drusenoid deposits. Gray level was measured once by a single grader, so the reproducibility of ROI placement was not assessed, and intraobserver agreement of pattern grading was not measured. Thirteen patients contributed both eyes; we accounted for this in the confidence intervals but not in the comparison among patterns. The ROI was fixed in size and centered on the fovea, so lesions outside the central 3.16 mm were not sampled. Finally, the independent gradings of the advanced AMD images were not retained in a form that allowed agreement to be computed, so advanced AMD is not reported.

## 5. Conclusions

Gray-level analysis of archived FAF images in early dry AMD is feasible with open-source software, but the resulting values are uncalibrated, did not differ significantly among Bindewald patterns in this cohort, and should not be compared between eyes or with quantitative autofluorescence. Independent grading of Bindewald patterns by two graders showed moderate agreement, lower than in the original description, and agreement on foveal involvement and lesion extent was fair. Studies and automated tools that rely on FAF pattern labels should report measured interobserver agreement and use explicit definitions and adjudicated labels.

## Data Availability

Data supporting the findings of this study are available from the corresponding author upon reasonable request.

## Author contributions

JNJ-H: conception and design, image analysis and pattern grading (first grader), data collection, analysis and interpretation, drafting of the manuscript. GG-S: pattern grading (second grader), data collection. AYH-V: review of discordant cases (third grader), critical revision, supervision. AS-C: supervision, administrative support. AB-S: supervision, critical revision. JAR-E: departmental supervision, critical revision for important intellectual content. All authors approved the final version and agree to be accountable for all aspects of the work.

## Ethics statement

The study was approved by the Institutional Ethics and Research Committees of Fundación Hospital “Nuestra Señora de la Luz,” I.A.P. and was conducted in accordance with the Declaration of Helsinki. Informed consent was not required because of the retrospective, noninterventional design.

## Data availability statement

De-identified data supporting the findings of this study are available from the corresponding author on reasonable request.

